# Noninvasive repeated sampling technique reveals specific respiratory cytokine signatures for electronic-cigarette and dual-product users obtained from a remote cohort of young adults

**DOI:** 10.1101/2025.10.23.25338090

**Authors:** Jennifer L. Thies, Hannah J. Appleseth, Naosuke Yamaguchi, Pria G. Bose, Adam M. Speen, Natalia Peraza, Catalina Cobos-Uribe, Alexia A. Perryman, Alexis A. Graham, Alayna P Tackett, Meghan E. Rebuli

**Author notes:** Corresponding author: Meghan E. Rebuli, PhD, 4310 Mary Ellen Jones Building CB 7325, Chapel Hill, NC 27599. **Conflict of Interest:** None to Report.

## Abstract

E-cigarette use has been linked to respiratory mucosal inflammation and other markers of toxicity. Dual use, or the use of e-cigarettes in combination with conventional cigarettes or other inhaled products has increased in prevalence, but there is limited understanding of the health effects associated with dual use. The aim of this study was to establish whether nasal mucosal cytokine profiles among never tobacco users, exclusive electronic cigarette users, and dual tobacco product users change over time and whether dual use significantly differs from exclusive e-cigarette use. This study utilized a repeated sampling study design, collecting nasal epithelial lining fluid from young adult participants (n=64) who were never tobacco users, exclusive electronic cigarette users, and dual tobacco product users, once weekly for four weeks using a remote, non-invasive sampling technique. Nasal mucosal immune mediators and salivary cotinine were then analyzed by ELISA. Differences in mucosal immune mediators were identified between e-cigarette users, dual tobacco product users and never users; however, these markers did not vary across time within group. E-cigarette and dual tobacco product users exhibited increased proinflammatory markers compared to never users. Chemokine profiles were uniquely altered in dual tobacco product users. Sex differences were identified in cytokine and chemokine production across groups.

These results suggest that remote, non-invasive nasal sampling is adequate for assessing immune profiles from tobacco product users and cross-sectional sampling is likely representative of consistent respiratory immune profiles across multiple weeks.

Dual product use results in distinct respiratory immune profiles, which suggests that long term disease outcomes may differ from exclusive product users.

**Implications:** This study demonstrates that respiratory mucosal immune mediator profiles altered with e-cigarette and other tobacco product use are stable over a period of weeks, suggesting that prior cross-sectional study results likely are representative of effects over longer periods of time. This study also shows that dual use of e-cigarettes with other tobacco products induces a unique elevated chemokine profile compared to sole e-cigarette use, while retaining similar elevated inflammatory cytokine profiles. This suggests that dual product use may induce differential long-term effects that sole product use.

## INTRODUCTION

Over the last decade, e-cigarette (EC) use has significantly increased, particularly in the young adult and adolescent population, raising a legitimate concern to the impact of usage on respiratory health and adverse side effects of use (1). In 2019, usage rates reached an all-time high among youth and young adult populations (2). The recommendation that ECs can be used as a safer alternative and smoking cessation tool in most countries has led to an increase in use (3,4), despite emerging evidence suggesting use, especially in combination with other products (dual use or EC+OTP), is associated with increased risk of long-term respiratory disease. Appealing flavorings, different types and brands, and targeted marketing make ECs more appealing to young adult and youth populations, sparking increased curiosity for use (5–7). Studies have shown the primary user group and consumers of ECs are young adults between 18-24 years of age, in the United States and other countries (8–12). Moreover, data indicates 40% of young adults that currently use ECs were never smokers before trying these devices, indicating this population is more vulnerable to marketing campaigns compared to other groups (13). Even devices that are labeled nicotine–free need to be approached with caution, as several studies have detected varying levels of nicotine within the tested e-liquid of these devices, misleading consumers(14–16).

Understanding the long-term consequences of EC use will still require more time and research. Despite years of study prior to 2019, a new consequence of EC use emerged in the form of EC or vaping product use-associated lung injury (EVALI), suggesting we have not identified all of the likely long-term consequences associated with such use (17–21). Even with the elimination of Vitamin E acetate in EC products, the likely contributor of this lung condition, and the decline of its presence on the market, there are still documented cases of EVALI (22). This suggests that illicit THC vapes, and cannabis use, may be a contributor. For example, an epidemiological study noted an increased risk of developing chronic obstructive pulmonary disease in a study of EC users, while dual product users displayed increased incidence and risk of all outcomes including cardiovascular disease, COPD and hypertension (23). Several components of ECs, including flavorings and other additives, have been linked to inflammation and other markers of toxicity, which may contribute to long-term disease (24–26). *In vitro* and *in vivo* studies suggest EC aerosols can activate immune cells, impairing key functions, elicit proinflammatory effect, impair viral clearance and cellular gene expression (27,28). Similar to conventional cigarettes, ECs increase the risk of cardiovascular disease, respiratory illness and immune system dysfunction, including increased inflammatory responses and susceptibility to infections (18,29,30). However, due to the constantly evolving EC market, additional studies are needed to address health effects of current product use patterns, particularly dual use.

Dual use, or the use of ECs in combination with conventional cigarettes or other inhaled products (tobacco, CBD, THC etc.) has increased in prevalence, particularly in the youth and young adult populations (31–34), reaching concerning levels (35). A 2019 population-based study by Xie and colleagues, suggested that more than 6 out of 10 EC users were dual users in the United States (34). There is limited understanding of the health effects associated with dual use, as it has not been as extensively studied as EC or conventional smoking have independently. However, the combination of inhaling combustible product smoke and ECs aerosols could lead to increased pathological outcomes compared to either alone. Additionally, there are reported increased risks for cardiovascular, respiratory, and immune system health (36).

To date, minimal longitudinal studies have been conducted to gather respiratory and immune outcomes of EC use. Likewise, with the emergence of dual tobacco use, limited data exists on the consequences of or outcomes of such exposure. To understand the comprehensive effects of EC and dual use, repeated-measure studies are necessary.

Previous studies on EC use are either primarily survey based with no biomarker confirmation or are cross-sectional and require participants to come into the clinic to provide samples to obtain respiratory biomarker data (23,37–39). Current literature frequently cites the limitation that it is uncertain whether observed biological effects are maintained with varying product use over longer periods of time due to lack of long-term follow-up studies (40–42). Moreover, these studies primarily focus on a few commonly measured pro-inflammatory immune mediators like IL-6, IL-8, IL-1β and TNF-α, potentially missing other respiratory immune consequences of use. To address this gap, the study herein fills an important and understudied research space.

We report one of the first repeated measure, longitudinal EC studies to be conducted remotely, using a novel non-invasive technique that allows for self-sampling and repeated collection of nasal epithelial lining fluid (NELF) biospecimens from e-cigarette users (EC), e-cigarette + other tobacco product users (EC+OTP), and never-users (CTRL), to evaluate a large cytokine profile for effects of use over four weeks. Overall, remote research methods are a feasible and easy way to obtain tobacco related respiratory health data from biospecimens across time (43). Results indicate immune mediator cytokine levels between all groups do not differ significantly week to week over the sampling period. They also demonstrate unique nasal mucosal immune mediator profiles in never users, EC users, and EC+OTP users, suggesting consistent and lasting respiratory effects of product use, despite variability in type of device or amount of use from week to week.

## MATERIALS AND METHODS

### Primary study design

Participants from the Biomedical Respiratory Effects Associated through Habitual Use of E-Cigarettes [BREATHE] study between 18 and 25 yrs of age were recruited for participation from the greater Los Angeles area, of Southern California, USA, between April – October 2021. Inclusion and exclusion criteria have been previously described (43), but briefly, participants were required to have a) ownership of a smartphone and willingness to download applications; b) current exclusive e-cigarette use for the past 3-months and reported never trying other tobacco products (EC), use of e-cigarettes and other tobacco products over the past 3-months (EC+OTP), or no use of e-cigarettes or tobacco products for the past 6-months (CTRL); c) ages 18-25; and d) fluency in English. Participants were excluded if they a) self-reported diagnosis of lung disease including cystic fibrosis, asthma, or chronic obstructive pulmonary disease b) had unstable or significant psychiatric conditions c) a history of cardiac event or distress within the past 3-months or d) currently pregnant or breastfeeding or planning to become pregnant (43). This protocol was reviewed and approved by the University of North Carolina – Chapel Hill, The Ohio State University, and The University of Southern California, Institutional Review Boards (IRB# 21-1077, 2023H0117, and UP-20-00992). After completion of the study and collection of diary logs, original enrollment group assignments were revisited. Cannabis was not included in the exclusion criteria.

However, after review, when participant logs indicated primarily cannabis use, these individuals were placed in a separate category for comparison (CAN).

### Sample Collection and Processing

Nasal epithelial lining fluid (NELF) was collected as previously described (43,44). Saliva samples were collected by collecting passive drool using standard methods. Participants were provided with instructions in their sampling kits for how to store samples until shipment. All sample specimens were stored in participants’ freezers until scheduled return pickup service was completed on days 8 and 22. Samples were collected 4 times, occurring once weekly during each study check-in visit conducted virtually to confirm the participant provided the sample accurately. All samples were stored at-80°C until shipment for processing. Samples were processed as previously described (44,45). As a nasal sampling strip was collected from both participant nares at each of the weekly sampling, a strip was randomly selected from either nostril for each participant at each visit to be used for downstream analyses. Prior studies suggested there is limited variability from nare to nare in individuals (44). NELF was eluted as described previously with 1% BSA with 0.05% Triton X-100 in Dulbecco’s phosphate-buffered saline (DPBS) and centrifugation. The resulting NELF eluate was stored at −20°C for analysis. Participants collected passive drool into a collection tube. Saliva was stored at –20°C until analysis.

### Saliva cotinine measurement

Cotinine, a nicotine metabolite, was used to evaluate exposure to nicotine-containing products. Cotinine was measured in saliva samples from each participants via ELISA per manufacturer instructions (Salivary Cotinine ELISA Kit, Salimetrics, Carlsbad, CA (Cat No: 1-2002)). For samples from EC or EC+OTP users, saliva was diluted by a factor of 10, while samples from CTRL were run with no dilution per manufacturer recommendation. For samples below the limit of detection for the assay, a value of zero was assigned.

### Protein extraction and measurement of cytokine levels

Soluble inflammatory mediators were analyzed with multiplex ELISA kit according to manufacturer instructions. A V-PLEX Human Cytokine 30-plex, consisting of V-PLEX Cytokine Panel 1, V-PLEX Chemokine Panel 1, and V-PLEX Proinflammatory Panel 1, were used (Mesoscale Discovery, Rockville, MD). Electrochemiluminescence was measured using a MESO QuickPlex SQ 120 (Mesoscale Discovery, Gaithersburg, MD). Final analysis yielded measurements across 29 total cytokines, with concentration values in pg/mL for each sample collected. Concentration values were adjusted for differences in processing and dilution volumes. Dilutions were conducted based on manufacturer recommended minimum dilutions for each kit.

### Data analysis

All analyses were conducted using R, v. 4.3.1using base R unless otherwise noted (R Core Team). Packages utilized for this manuscript include *dplyr* (46), *tidyr* (47), *patchwork* (48), *ggplot2* (49), *ggprism* (50), *magrittr*, (51) *ggpubr* (52), *rstatix* (53), *formattable* (54), and *ggfortify* (55). Participants who did not have a complete set of data from all four visits were removed from the final data set (n=13, Supplementary Figure 1). Missingness based on values below the assay limit of detection was handled by imputation. The value imputed was determined by taking the minimum value for each particular analyte and dividing by 2 (56,57).

Normality was tested using a Shapiro-Wilk test. Non-parametric tests were used for all comparisons. For group cytokine differences, sex differences, and visit differences, a Kruskal-Wallis test, followed by a Dunn’s test for nonparametric multiple comparisons was used on raw data for each cytokine within each user group. Because there were no differences between visits, analyses used combined data. For cotinine analyses, a Kruskal-Wallis test was also performed. Correlation analyses were conducted for each analyte compared to cotinine concentrations within the specified groups using the Spearman’s method.

## RESULTS

### Participant demographics

Overall, 64 participants were enrolled in the primary cohort of the study. Demographic descriptions are summarized for all participants in **Table 1**. Among the individuals included in the study (n = 64), 14 were e-cigarette exclusive users (EC), 10 were e-cigarette + other tobacco product users (EC+OTP), 6 were cannabis users (CAN), and 34 were never users (CTRL). To be enrolled in the study participants needed to be a non-tobacco user, EC user, or EC+OTP user. CAN participants originally specified themselves as non-tobacco users but were later identified as cannabis users throughout the study based on diary entries provided.

**Table 1:**
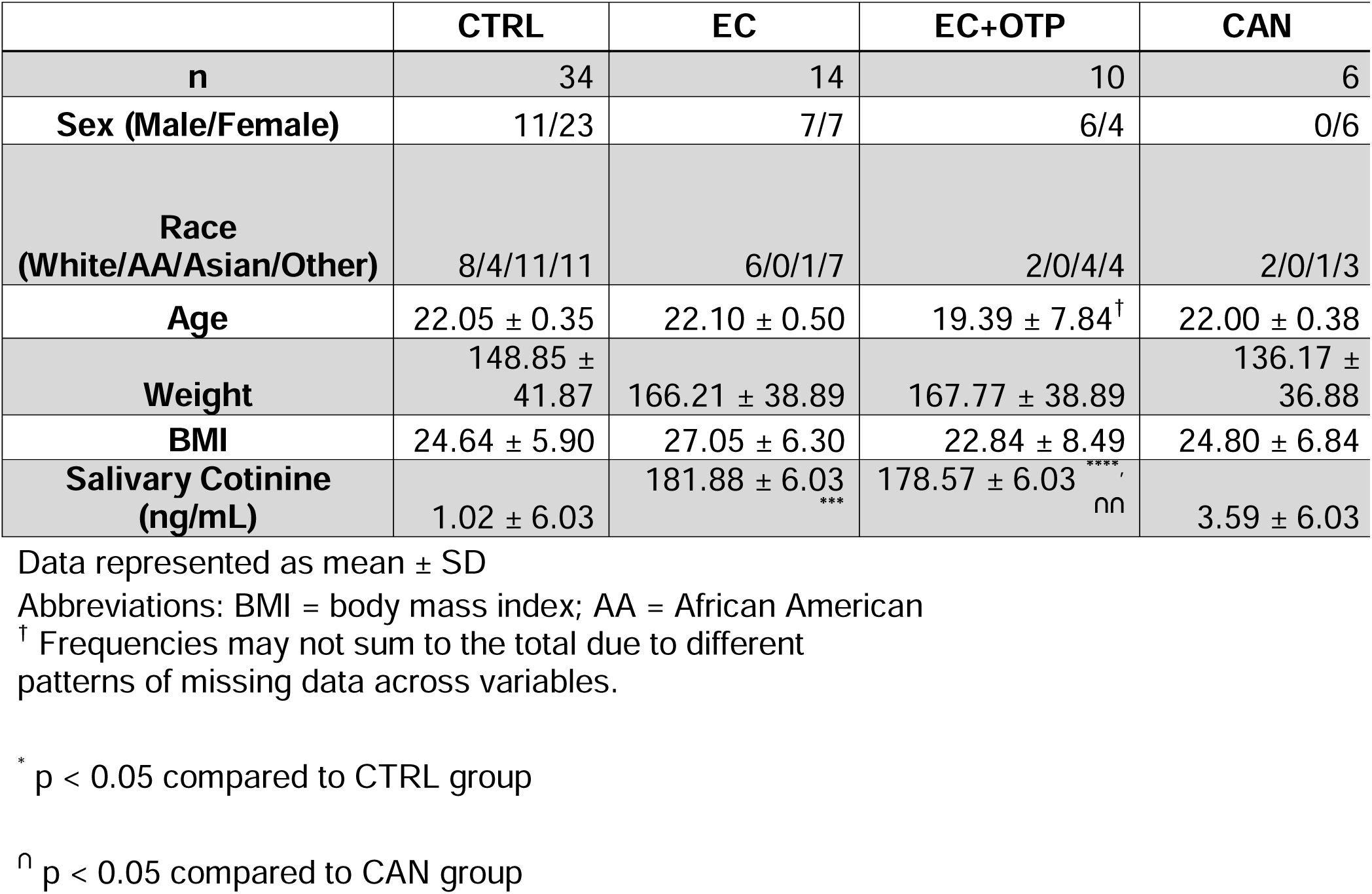
Participant Demographic Information.

|  | **CTRL** | **EC** | **EC+OTP** | **CAN** |
| --- | --- | --- | --- | --- |
| **n** | 34 | 14 | 10 | 6 |
| **Sex (Male/Female)** | 11/23 | 7/7 | 6/4 | 0/6 |
| **Race**<br>**(White/AA/Asian/Other)** | 8/4/11/11 | 6/0/1/7 | 2/0/4/4 | 2/0/1/3 |
| **Age** | 22.05 ± 0.35 | 22.10 ± 0.50 | 19.39 ± 7.84<sup>†</sup> | 22.00 ± 0.38 |
| **Weight** | 148.85 ± 41.87 | 166.21 ± 38.89 | 167.77 ± 38.89 | 136.17 ± 36.88 |
| **BMI** | 24.64 ± 5.90 | 27.05 ± 6.30 | 22.84 ± 8.49 | 24.80 ± 6.84 |
| **Salivary Cotinine**<br>**(ng/mL)** | 1.02 ± 6.03 | 181.88 ± 6.03<sup>***</sup> | 178.57 ± 6.03<sup>****, nn</sup> | 3.59 ± 6.03 |

Data represented as mean ± SD
Abbreviations: BMI = body mass index; AA = African American
<sup>†</sup> Frequencies may not sum to the total due to different patterns of missing data across variables.
\* p < 0.05 compared to CTRL group
<sup>nn</sup> p < 0.05 compared to CAN group

Because cannabis use was not part of the exclusion criteria, these subjects were moved into their own grouping category for the analyses within this study. Of the 64 subjects, 62.5% (n=40) were females and 37.5% (n=24) were males. Most participants identified their sexual orientation as straight, with 20% of individuals identifying as either (gay, bisexual, pansexual or asexual). The distribution of race and ethnicity among participants was consistent, with no significant differences identified between user groups. The average age of the entire cohort was 21.7 (range of 21.5-23.2) years. The average age for CTRL, EC users, EC+OTP users and CAN users, was 22.05, 22.10, 19.39 and 22.00 years, respectively. The overall average BMI was 24.83 (range of 16.40 - 44.90). Lastly, the average weight was 154.75 (range of 86-270) lbs. These characteristics were consistent across user status, as no significant differences were identified for age, weight, or BMI. Analyses were conducted as aggregate group analyses by EC, EC+OTP, CAN, and CTRL, and with consideration of sex as a biological factor in all groups except for CAN users, where the N was insufficient for sex stratified analyses.

### Baseline measurements

#### Cotinine levels

Cotinine levels collected from nasal epithelial lining fluid (NELF) across all subjects had an average of 63.12 (range of 0-1750.51) ng/mL. By group, CTRL had average cotinine level of 1.02 (ranging from 0-5.58) ng/mL. Comparatively, EC users which had an average cotinine concentration of 181.88 (range 0-1750.5) ng/mL, EC+OTP had an average of 178.57 (ranging from 0-930.83) ng/mL and CAN users with an average of 3.59 (ranging from 0-23.57) ng/mL. Nicotine concentrations were greater in EC users vs. CTRL and EC+OTP vs. CTRL (*p* < 0.05). Interestingly, nicotine concentrations were elevated in EC+OTP user compared to CAN users (*p* < 0.05) (**Figure 1**). Elevated cotinine levels in both EC and EC+OTP users were expected, as these devices contain nicotine (38). No sex differences were identified in cotinine levels, even when separated by group.

**Figure 1:**
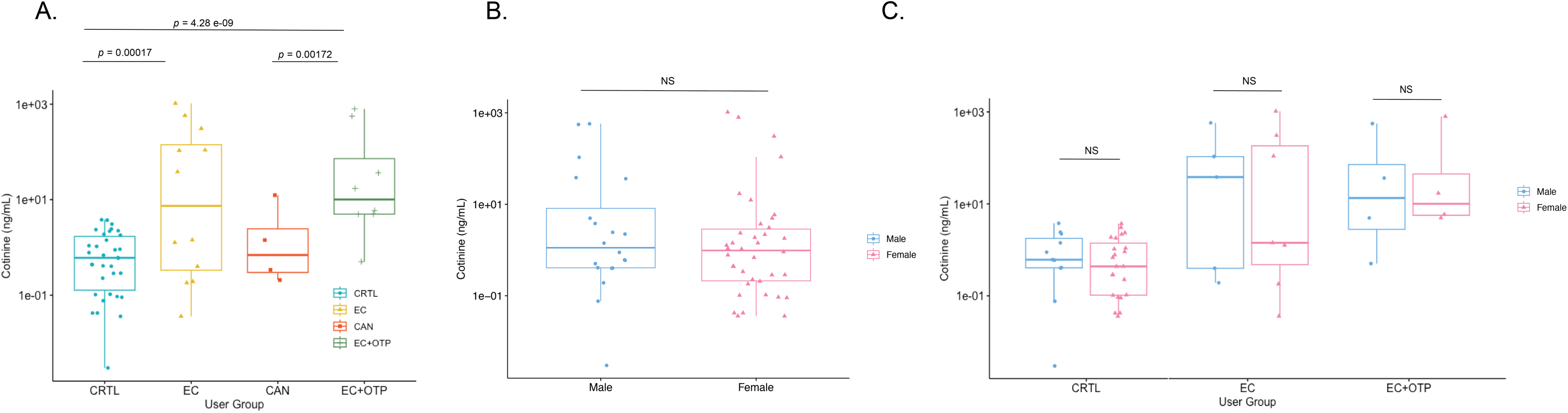
Salivary cotinine differences between user groups and by sex. Each point represents the average of 4 visits per individual. Center line within boxes represents median cotinine concentration within each group. (A) Salivary cotinine levels were significantly elevated for EC and EC+OTP users compared to CTRL. EC+OTP users also displayed increased cotinine compared to CAN users. (B) No significant differences were identified when cotinine levels were analyzed by sex in aggregate or (C) when separated by group. Statistics are represented with exact p-values. Differences between exposure groups and sex were identified using a Kruskal-Wallis test. If results were significant, a Dunn’s *post hoc* test was used to follow up on significant differences.

#### General cytokine distributions

When evaluating immune mediator analytes that displayed the greatest percent change compared to CTRL, distributions varied between user <u>groups</u> (**Supplementary Table 1**). Analytes IL-2, IL-17, and MIP-1β exhibited the greatest percent increase for EC users compared to CTRL. Analytes IL-5, MIP-1β and Eotaxin-3, displayed the greatest increase for CAN users, and IL-5, Eotaxin-3 and MIP-1β displayed the greatest increase for EC+OTP users. Interestingly, MIP-1β was among the greatest changed analyte between all groups. The same analytes displayed the greatest changes for the CAN and EC+OTP user groups, albeit in different orders, possibly suggesting more similarities in their signatures. Both EC and EC+OTP user groups saw positive increases in all tested analytes, indicating increased concentration levels compared to CTRL. Within the CAN group, half the analytes displayed a percent change decrease, indicating lower immune mediator concentration levels compared to CTRL.

##### Immune mediators do not differ over sampling period

NELF samples were taken by participants once weekly over the four week study period. To identify whether immune mediators changed across time, we analyzed participants by visit. No differences were identified across time for any of the tested analytes, regardless of user status (**Figure 2**). This suggests soluble protein mediators remain stable over time.

**Figure 2.**
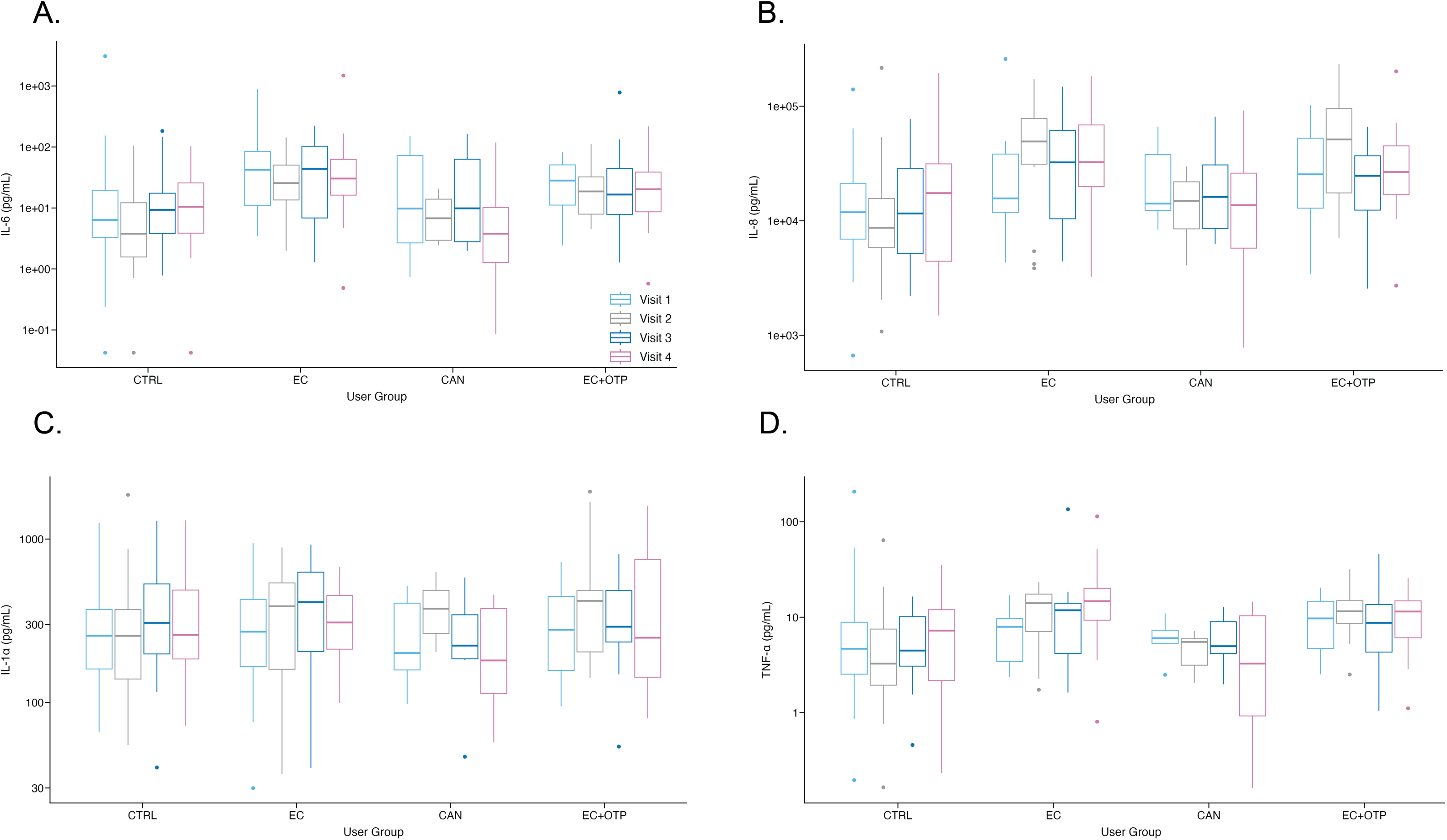
Visit differences across each user group. No visit differences were identified in any users group across time for any analyte. Representative analytes shown here are (A) IL-6 (B) IL-8 (C) IL-1α and (D) TNF-α.

##### EC and EC+OTP use increase proinflammatory cytokines

To further understand the respiratory immune differences induced by EC use, we carried out statistical analyses at the individual immune mediator level for 29 cytokines and chemokines. As there were no detected differences by time across the four weeks of sampling (**Figure 2**), we utilized an average value across weeks for evaluation of user group differences in biomarker levels. We used a Kruskal-Wallis test followed by a Dunn’s test to identify differences between groups. Overall, mean concentrations of 19 analytes were found to be significantly different (*p* < 0.05) between CTRL and EC users and CTRL and EC+OTP users, many of which are Th1 and Th2 cytokines: GM-CSF, IL-12p40, IL-16, IL-17, IL-5, TNF-β, VEGF, Eotaxin-3, IL-8, MDC, MIP-1α, MIP-1β, IL-10, IL-13, IL-1β, IL-2, IL-4, IL-6, TNF-α (**Figure 3A-D, Supplementary Figure 2 and Supplementary Table 2**). Independently, two additional analytes, IFN-γ and IL-12p70, were only significantly different from EC users to CTRL. EC+OTP users displayed a unique signature independent of other cohorts. Participants exhibited an increase in four chemokines Eotaxin, MCP-1, TARC and MCP-4 (*p* < 0.05) compared to CTRL (**Figure 3E-H**). These data demonstrate EC and EC+OTP use, increases production of proinflammatory immune mediators in the nasal mucosa, with EC+OTP use uniquely elevating additional nasal mucosal chemokine levels.

**Figure 3:**
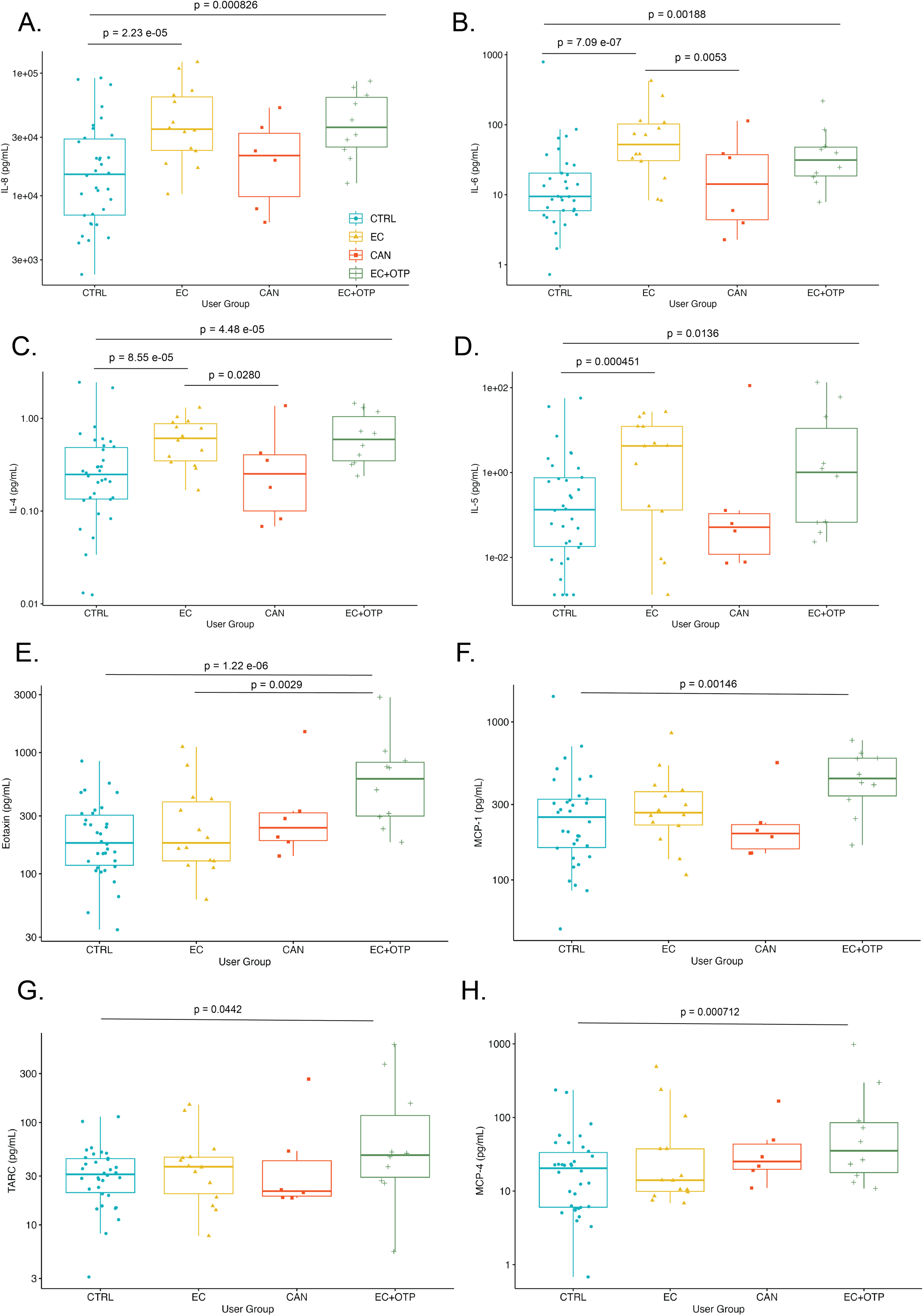
EC and EC+OTP users displayed increases in proinflammatory soluble mediators compared to CTRL, but only EC+OTP users showed increased specific chemokine levels. Select proinflammatory cytokines that were significantly different for EC users and EC+OTP users compared to CTRL including: (A) IL-8 (B) IL-6 (C) IL-4 and (D) IL-5. Chemokines (E) Eotaxin (F) MCP-1 (G) TARC and (H) MCP-4 display significantly elevated protein levels in EC+OTP users only compared to CTRL. Each point represents the average of 4 visits per individual. Center line within boxes represents median protein concentration within each group. Statistics are represented with exact p-values, and differences between exposure groups were identified using a Kruskal-Wallis test followed by Dunn’s *post hoc* test.

##### Cannabis differences compared to EC and EC+OTP users

Within the CAN user group, participants did not display any significant differences compared to CTRL; however, differences compared to the EC and EC+OTP users were apparent. For 15 analytes: GM-CSF, IL-12p40, IL-16, IL-17, TNF-β, MCP-1, MIP-1β, IFN-γ, IL-10, IL-12p70, IL-13, IL-1β, IL-2, IL-4, IL-6, and TNF-α, cannabis use alone displayed decreased expression compared to EC and EC+OTP use (*p* < 0.05). Ten of the analytes displayed lower concentrations for the CAN group compared to the EC+OTP user group and twelve analytes were lower for the CAN group compared to the EC group. Six analytes displayed lower concentrations for CAN users for both groups (**Figure 4 and Supplementary Figure 3 and Supplementary Table 2**). These data suggest that cannabis use does not affect the measured respiratory immune mediators in the same way as EC or EC+OTP usage.

**Figure 4:**
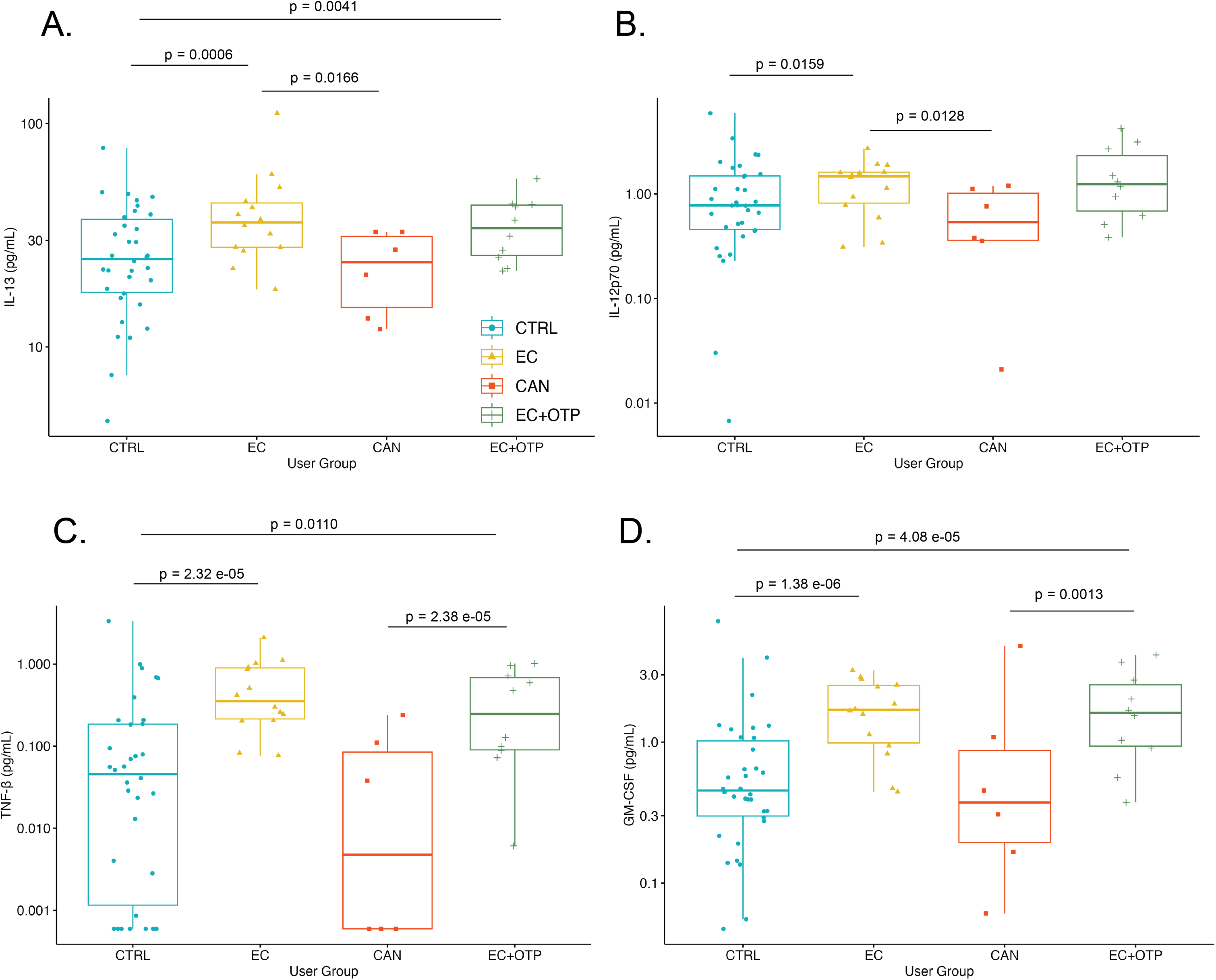
CAN users display lower concentrations of soluble mediators compared to EC and EC+OTP users. Selected immune mediates that are lower in CAN users include: (A) IL-13 and (B) IL-12 present significantly lower concentrations in CAN users compared to EC users. (C) TNF-b and (D) GM-CSF are significantly lower for CAN users compared to EC+OTP users. Differences between exposure groups were identified using a Kruskal-Wallis test followed by Dunn’s *post hoc* test. Each point represents the average of 4 visits per individual. Center line within boxes represents median protein concentration within each group.

##### Analytes with no effect

While for a majority of the measured 29 immune mediators differences were identified in at least one user group, some analytes displayed no differences (IL-15, IL-1α, IL-7 and IP-10). This suggests their role in immune function is not altered between user groups (**Supplementary Figure 4)**.

##### Sex differences in analytes

As prior studies have shown sex specific effects of EC use (58–60),we further stratified the data by testing whether individual analytes associated with each user group displayed differences between sexes and whether it was a main effect of sex or an interaction effect. Of the 29 tested analytes, 24 had main effect sex differences: GM-CSF, IL-12p40, IL-16, IL-17, IL-1α, IL-5, IL-7, TNF-β, VEGF, Eotaxin-3, IL-8, IP-10, MDC, MIP-1α, MIP-1β, IFN-y, IL-10, IL-12p70, IL-13, IL-1β, IL-2, IL-4, IL-6, and TNF-α (*p* < 0.05) while 17 analytes displayed group by sex interactions: GM-CSF, IL-16, IL-17, IL-1α, IL-5, IL-7, TNF-β, VEGF, IL-8, IP-10, MIP-1β, IL-10, IL-12p70, IL-1β, IL-2, IL-4, and TNF-α (*p* < 0.05) (**Supplementary Table 3**). Due to the low number of participants within the CAN group, and the group only containing females, this group was excluded from sex difference results. For most of the analytes that displayed group by sex interactions, in post-hoc testing, males displayed higher concentrations compared to females <u>except</u> for IL-7 and IP-10 in the CTRL and IL-10 for the EC+OTP users. There were several other patterns of sex-specific effects observed and described in more detail below, elimination of non-user detected sex differences with exposure, introduction of exposure-specific sex differences when none existed in the non-user group, and changes in magnitude of sex differences by user group.

##### Elimination of sex differences with product use

In several analytes significant sex differences were apparent in the nonuser group but lost in either the EC group, EC+OTP user group or both. Analytes GM-CSF, TNF-β, IL-1α, and IL-2 lost sex differences in the EC group only, while differences remained in the EC+OTP user group. Mediators IL-5, IL-7, IP-10, IL-12p70 and IL-4, lost sex differences in both EC and EC+OTP user groups (**Figure 5A-D and Supplementary Figure 5**). Worth noting, IL-7 displayed baseline sex differences where females exhibited higher concentration levels. When user status was considered, sex differences were no longer apparent in both the EC and EC+OTP user groups. Likewise, IP-10 displayed significant baseline sex differences where females showed elevated concentrations. Sex differences were eliminated when user status was considered in both EC and EC+OTP user groups, with females trending lower than males. IL-7 and IP-10 were the only cases where analyte levels were significantly increased in females compared to males.

**Figure 5:**
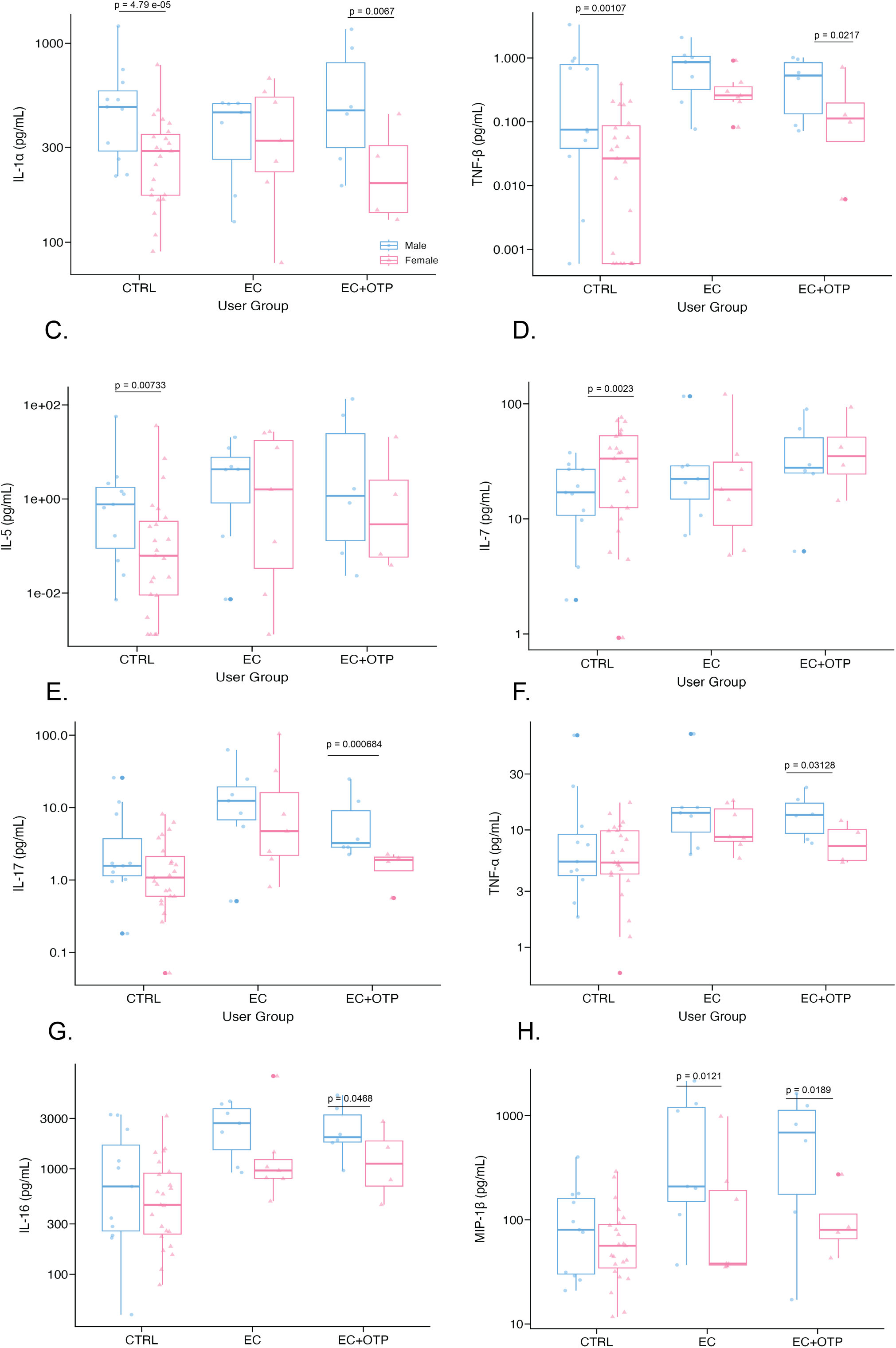
Changes in sex differences by product use group. Baseline sex differences are lost in some immune mediators depending on user status. Nonuser sex differences that were identified in (A) IL-1α and (B) TNF-β were lost in the EC user group specifically. Nonuser sex differences that were identified in (C) IL-5 and (D) IL-7 were lost in both EC and EC+OTP user groups. User status can also influence sex differences not seen in the CTRL group, but that are introduced with product use. Baseline sex differences in CTRL subjects were not apparent for (E) IL-17 (F) TNF-α or (G) IL-16, however, EC+OTP users displayed significant sex differences and for (H) MIP-1β sex differences were identified in both EC and EC+OTP users. Each point represents the average of 4 visits per individual. Center line within boxes represents median analyte concentration within each group. Statistics are presented with exact p-values. Differences between sex within each exposure groups were identified using a Kruskal-Wallis test followed by Dunn’s *post hoc* test.

##### Introduction of sex differences with product use

For analytes IL-16, IL-17, MIP-1β, IL-10, TNF-α, and IL-8, no sex differences were identified in the nonuser group, however, when taking EC or EC+OTP use status into consideration, sex differences became apparent in one or both groups (**Figure 5E-H and Supplementary Figure 6**). IL-8 gained sex differences in the EC group, after no sex differences were detected in CTRL. The EC+OTP user group trended towards a sex difference, although it was not statistically significant (*p* = 0.0644). The magnitude in which IL-8 was detected did increase in both user groups. IL-16 and MIP-1β acquired sex differences in both user groups. The magnitude in which these analytes were expressed was increased as well. IL-17, IL-10, and TNF-α gained sex differences in the EC+OTP user group. Interestingly, IL-10, which did not display baseline sex differences, presented with sex differences in the EC+OTP user group. When considering user status, females displayed higher IL-10 concentration levels, something that was not seen in other analytes. VEGF trended towards sex differences in the EC+OTP user group, albeit not significantly (*p* = 0.0644).

##### Amplified sex differences with product use

Also worth noting is that many analytes displayed changes in magnitude as a response to EC or EC+OTP use (**Supplementary Figure 7**). IL-1β displayed sex differences in the non-user group. These sex differences were still visible in the EC group and the EC+OTP user group, albeit at a greater magnitude. A similar trend was seen with IL-16 and MIP-1β. Lastly, sex differences for TNF-β concentrations were amplified in the EC and EC+OTP user groups compared to the nonuser group. While the concentrations were not statistically significant between sexes in the EC group, they were in the EC+OTP user group, approximately 10 times greater.

##### Cotinine Associations

We also examined whether immune mediator concentrations obtained from NELF correlated with cotinine concentration levels in saliva to evaluate the role of nicotine in the observed responses. Minimal correlations were identified for EC users, CAN users, and CTRL. Analytes for CTRL that displayed significant correlations between cotinine and immune mediator concentrations included IL-6, IL-16, and IL-17; all were negative. For CAN users TNF-α, IL-13, and IL-8, indicated negative correlation between cotinine and protein concentrations. Similarly in EC users, TNF-α and IL-17 displayed significant values that were negatively correlative. Interestingly, EC+OTP users displayed the most correlations between analytes and cotinine levels.

14 analytes were negatively correlated (IL-4, TARC, MIP-1α, MIP-1β, MCD, MCP-1, MCP-4, IP-10, Eotaxin, Eotaxin-3, IL-5, IL-15, IL-16, and GM-CSF) (Supplementary Figure 8 **and Supplementary Table 4**).

We further stratified cotinine data within each group by sex, excluding the CAN group due to low N and a female sample bias. In general, males were the primary drivers of correlative patterns between the two sexes for the remaining three groups, CTRL, EC and EC+OTP. For CTRL females, only two analytes, IL-7 and TARC, displayed a significant, but positive, correlation with cotinine. Males displayed significant negative correlations with cotinine for 20 of the 29 analytes, TNF-α, IL-6, IL-4, IL-2, IL-13, IL-10, TARC, MIP-1β, MDC, MCP-4, IP-10, Eotaxin, Eotaxin-3, VEGF, IL-7, IL-5, IL-15, IL-16, IL-17, and GM-CSF. As for the EC users, females displayed negative correlations for TNF-α, MIP-1β, MCP-1, IL-5, and IL-17, while males displayed a positive correlations for MCP-4 and Eotaxin-3. For EC+OTP users, significant correlations were only identified in male users. 12 analytes, TARC, MIP-1α, MIP-1β, MDC, MCP-4, Eotaxin, Eotaxin-3, IL-4, IL-5, IL-15, IL-16, and GM-CSF, displayed negative correlations with cotinine (**Supplementary Table 5**).

## DISCUSSION

In this study, we aimed to evaluate cytokine signatures across different user groups including EC users, EC+OTP users, CAN users and CTRL using weekly repeated sampling over one month. Moreover, we sought to assess whether remote non-invasive sampling was feasible for obtaining biospecimens from a more diverse cohort of participants. These findings are imperative as they provide considerable evidence to support the effectiveness of remote, noninvasive sample collection and longitudinal studies. Several main findings were identified. First, analyses identified a significant increase in proinflammatory immune mediators in EC and EC+OTP use individuals compared to CTRL. Second, EC+OTP users show a specific chemokine signature apparent only with this category. Next, CAN user profiles did not consistently differ significantly compared to CTRL; however, compared to EC and EC+OTP users, CAN use displayed significantly lower measurements of cytokines. Further, sex specific differences in NELF immune mediators depend on user status. Lastly, nasal mucosal cytokine levels did not vary across the one month sampling period in any groups.

It is known that inflammatory markers are elevated among tobacco smokers (61,62), and it has been previously documented that EC users display increased levels of inflammatory biomarkers compared to CTRL but decreased compared to smokers (37). Prior *in vitro*, *ex vivo* and human studies have demonstrated EC aerosol exposure increases inflammatory cytokine profiles and alters immune function (21,63) including in mouse bronchial alveolar lavage fluid (BALF) where EC exposure increases Th1 and Th2 cytokines, proinflammatory cytokines and chemokines (60,64). Similarly, here we showed increases in levels of 21 different cytokines in EC users compared to CTRL including Th1, Th2 and proinflammatory cytokines. Thus, our findings of increased respiratory inflammation with EC use are consistent with field findings for third generation device use (38,65).

Minimal comparative analyses exist that evaluate and discuss the respiratory immune marker signature of EC+OTP use and the differences from exclusive EC use. To our knowledge, this is one of the first studies to assess the effects of EC+OTP use on immune mediators with repeated sampling. EC+OTP user participants displayed an increase in 23 of the tested cytokines compared to CTRL. Available data in the literature does suggest that EC+OTP use may induce increased health effects over exclusive EC use, including increased oxidative stress and inflammatory biomarkers (66), increased hs-CRP, a marker for cardiovascular disease, (67), and increased biomarkers of tobacco smoke and nicotine exposure (68). This is additionally supported by emerging *in vitro* studies (69). Furthermore, numerous vaping devices can now be used to inhale cannabinoids, an area in which limited knowledge exists, and data on the health effects of inhalation are not well understood. Although, few studies have indicated that vaping cannabinoids induced more potent inflammatory responses and greater oxidative stress than vaping nicotine alone (70,71). Overall, while we show evidence of distinct patterns of respiratory immune cytokine production with EC+OTP product use vs. CTRL and exclusive EC users, more research is needed to uncover the larger respiratory impact of EC+OTP use in comparison to effects of EC use, which is more well-studied (72).

Largely, CAN users displayed minimal differences in measured biomarkers compared to CTRL. However, significant differences were observed compared to EC and EC+OTP users. Minimal studies have evaluated the impacts of cannabis use on respiratory and airway immune biomarkers due to its status as a controlled substance, leaving the impacts of cannabis use poorly understood. One study observed similar airway inflammation and tissue damage in cannabis smokers compared to tobacco smokers, although their sample size was small (73). These results differ from what was found in our study, showing minimal change between CAN users and CTRL, although the N of CAN users in our study was also very small with a female sample bias and results should be interpreted as preliminary. Thus, discordance with the prior study is likely a function of low power. In mouse models of acute lung injury, cannabinoids have displayed anti-inflammatory properties in bronchoalveolar lavage (74). Cannabis constituents have also shown reduction in both acute and chronic inflammatory responses in rat when administered orally (75). Overall, the literature around the respiratory immune effects of cannabis use is sparce and larger prospective studies are needed to definitively evaluate health effects We identified several differences in inflammatory cytokines between sexes of all users groups, addressing a common limitation of other tobacco and EC studies by including sex-stratified analyses (76). Overall, 24 of the tested analytes displayed main effect sex differences and 17 analytes displayed group by sex interactions. Several sex-difference patterns were identified, including groups of analytes where baseline sex differences were eliminated due to product use, sex differences that were introduced when user status was considered, and sex differences that were present at baseline and amplified with product use. While most analytes displayed higher concentrations in males, IL-7, IP-10 and IL-10 exhibited greater concentrations in females. Additional sex-specific responses were identified in cotinine analyses as well. Identifying sex specific responses aligns with a variety of studies in the literature which have demonstrated sex specific effects of device and use preferences, biological response to EC ingredients, and effects of use on lung disease (77,78). Prior studies have also examined nasal cytokines in exclusive EC users in combination with models of viral infection and found sex by exposure interactions(45,79). These data further corroborate the importance of including sex when designing and completing studies and analyses. It remains unclear as to whether the sex differences observed are derived from sex specific product use, sex-differences in response to similar toxicant exposures, or a combination thereof.

We were interested in determining whether changes in immune mediators occurred over time with variability in use rate or product use. Many prior studies have been cross-sectional in nature and frequently cite a lack of longitudinal assessment as a limitation. Thus, we sought to address this gap in the literature. Participants collected four total samples, one sample per week, over the observational period. We tested each analyte for differences across time and observed no significant changes from week to week (**Figure 2**). These data indicate that despite some variability in product use rates and type of products used over time, that biological effects of EC and EC+OTP use result in consistent changes in respiratory immune mediator profiles. These findings also suggest that results from previous cross-sectional studies with a limited number of visits are (80–82) likely an accurate representation of immune mediator levels for users over time.

We measured salivary cotinine concentrations, a metabolite of nicotine, to better understand nicotine exposure in participants by group. In aggregate, limited correlations with immune mediators from NELF were identified for all user groups. Although no significant differences were identified between EC users and EC+OTP user groups in cotinine concentration, EC+OTP users displayed significant correlative values with 14 analytes to varying degrees of strength, suggesting the addition of other products creates a different immune environment and could be driving the additional immune mediator changes observed. Nicotine does not seem to be a primary driver of the observed cytokine changes by user group. Prior studies have suggested both the humectant and flavoring components may alter respiratory immune profiles (60) and could explain these differences. Likewise, different generation devices influence airway immune biomarkers differently and generate varying concentrations of nicotine and reactive oxygen species based on device mechanism (65). The male-driven correlations within each group where sex differences were considered, may be due to sex-specific differences in product use patterns, whereby sex or gender have been previously associated with type of EC, flavor, and nicotine dose use, as well as place of purchase (77,78,83). Variations in products used likely results in different chemical exposure profiles, contributing to varying immune mediator production by sex. Increased correlation with cotinine observed in males suggests more nicotine-dependent effects compared to females.

While our study provides novel insight into the differences of respiratory immune mediators across different user groups, limitations are important to consider. Sampling a larger cohort would allow for greater study quality and more power when running statistical tests – specifically when considering the CAN user group. The authors recognize results are generalized to a specific area in California. Additional studies evaluating sampling at a national level would be informative. Overall, our study included relatively small sample sizes, which limited the power to detect differences between groups and sex. Secondly, this study primary focused on a narrow age group of individuals. While young adults between the ages 18-24 are one of the highest users of ECs and other products, these data may not accurately represent use effects in older populations. Research has shown that EC generation and flavorings can impact immune responses (69). Separating participants based on device type or including device type information into exclusion criteria should be considered for future studies.

Differences in products could be a contributor to some of the intra-group variation seen in immune mediators. Furthermore, understanding more about the products (i.e., tobacco containing, cannabis, conventional) EC+OTP use participants used would be important to consider for further studies. Lastly, we recognize that some of our non-user participants were exposed to secondhand smoke.

## Conclusion

To our knowledge, this study was one of the first to evaluate the variability of nasal cytokine signatures across different user groups in a longitudinal study, as well as compare profile differences between EC and EC+OTP users. The results provide evidence that both EC and EC+OTP use increase nasal cytokine levels, that cytokine levels do not vary substantially from week to week, and that profiles between user groups are distinct. Further, we show that EC use, and EC+OTP use can affect the expression of sex differences in nasal immune mediator profiles. Overall, this work highlights that EC and EC+OTP use can alter biomarkers of respiratory function and suggests that prolonged use may result in long-term health effects. Furthermore, the use of noninvasive techniques such as NELF collection for biomarker analysis in the airways in remote settings, should be given greater consideration to provide increased opportunities for sociodemographic and socioeconomic diversity in study groups and health creating more equitable chances to advance scientific research.

## Author Contributions

APT conceived of and designed the study and obtained funding; APT, NY, NP, and MER contributed to sample and subject data collection; JLT, PGB, AMS, CCU, ANP, and MER analyzed samples; JLT, HJA, AAG, and MER analyzed data and created visualizations; all authors contributed to data interpretation; JLT and MER drafted the manuscript; all authors contributed to editing the manuscript and approve of the final version.

## Data Availability

All data produced in the present study are available upon reasonable request to the authors

## Acknowledgements

We would like to thank Jessica Barrington-Trimis and the research participants for their contributions to this study.

## Supplementary Figure Legends

**Figure S1:** Consort Diagram

**Figure S2.** Significant protein differences between EC and EC+OTP user groups compared to CTRL. (A) Eotaxin-3, (B) GM-CSF, (C) IL-1b, (D) IL-2, (E) IL-10, (F) IL-12p40 (G) IL-12p70 (H) IL-13, (I) IL-16 (J) IL-17 (K) MCD (L) MIP-1α (M) MIP-1β (N) TNF-α (O) TNF-β and (P) VEGF displayed increased protein concentrations (pg/mL) compared to CTRL.

**Figure S3.** Proteins that are significantly lower in cannabis users compared to EC and EC+OTP. (A) IFN-γ, (B) IL-1β, (C) IL-2, (D) IL-10, (E) IL-6, (F) IL-12p40 (G) IL-16 (H) IL-17, (I) MCP-1 (J) MIP-1β and (K) TNF-α displayed decreased protein concentrations (pg/mL) compared to EC and EC+OTP use users.

**Figure S4.** Soluble proteins with no significant differences across users groups. (A) IL-15, (B) IL-7, (C) IP-10 and (D) IL-1α.

**Figure S5.** Baseline sex differences that were lost based on user status. Baseline sex differences were observed in (A) IP-10 (B) IL-12 (C) IL-2 and (D) IL-4, but lost in the EC and/or EC+OTP use user groups

**Figure S6.** Analytes with no baseline sex differences, but sex differences observed in user groups. (A) Sex differences were introduced for IL-8 in the EC user group and (B) IL-10 had sex differences introduced for the EC and EC+OTP user groups.

**Figure S7.** Sex differences with magnitude changes (A) IL-1β (B) TNF-β (C) IL-16 and (D) MIP-1β

**Figure S8:** Salivary cotinine and protein data correlations. Correlation of cotinine concentrations (y-axis) and immune protein concentrations (x-axis) for all user groups. (A) CTRL IL-17 (B) EC users TNF-α (C) CAN users IL-8 and (D) EC+OTP users TARC.

## Supplementary Table Legends

**Table S1**: Percent Change for Analytes across Group compared to CTRL

**Table S2:** Aggregate group differences for each analyte and the associated significance between each group by analyte.

**Table S3**. Effects of sex as an interaction and main effect for user groups and their associated significance by analyte

**Table S4**. Salivary cotinine correlations for each user group by analyte. Bolded values indicate significant correlations between cotinine and protein concentrations. Color indicates strength of correlation.

**Table S5**. Salivary cotinine correlations for each user group separated by sex and analyte. Bolded values indicate significant correlations between cotinine and protein concentrations. Color indicates strength of correlation.

