## Supplementary Figures for "Noninvasive repeated sampling technique reveals specific respiratory cytokine signatures for electronic-cigarette and dual-product users obtained from a remote cohort of young adults"

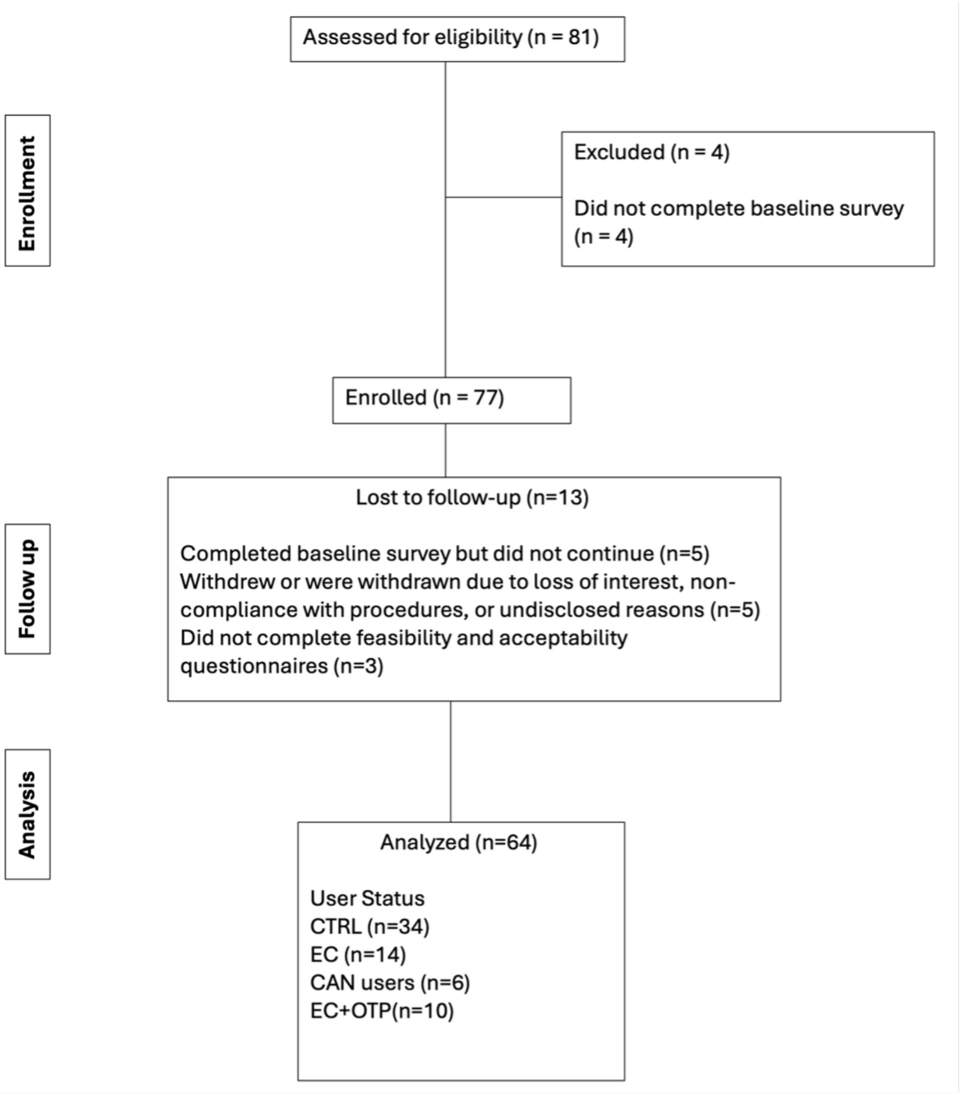


**Supplementary Figure 1**

### Supplementary Figure 2

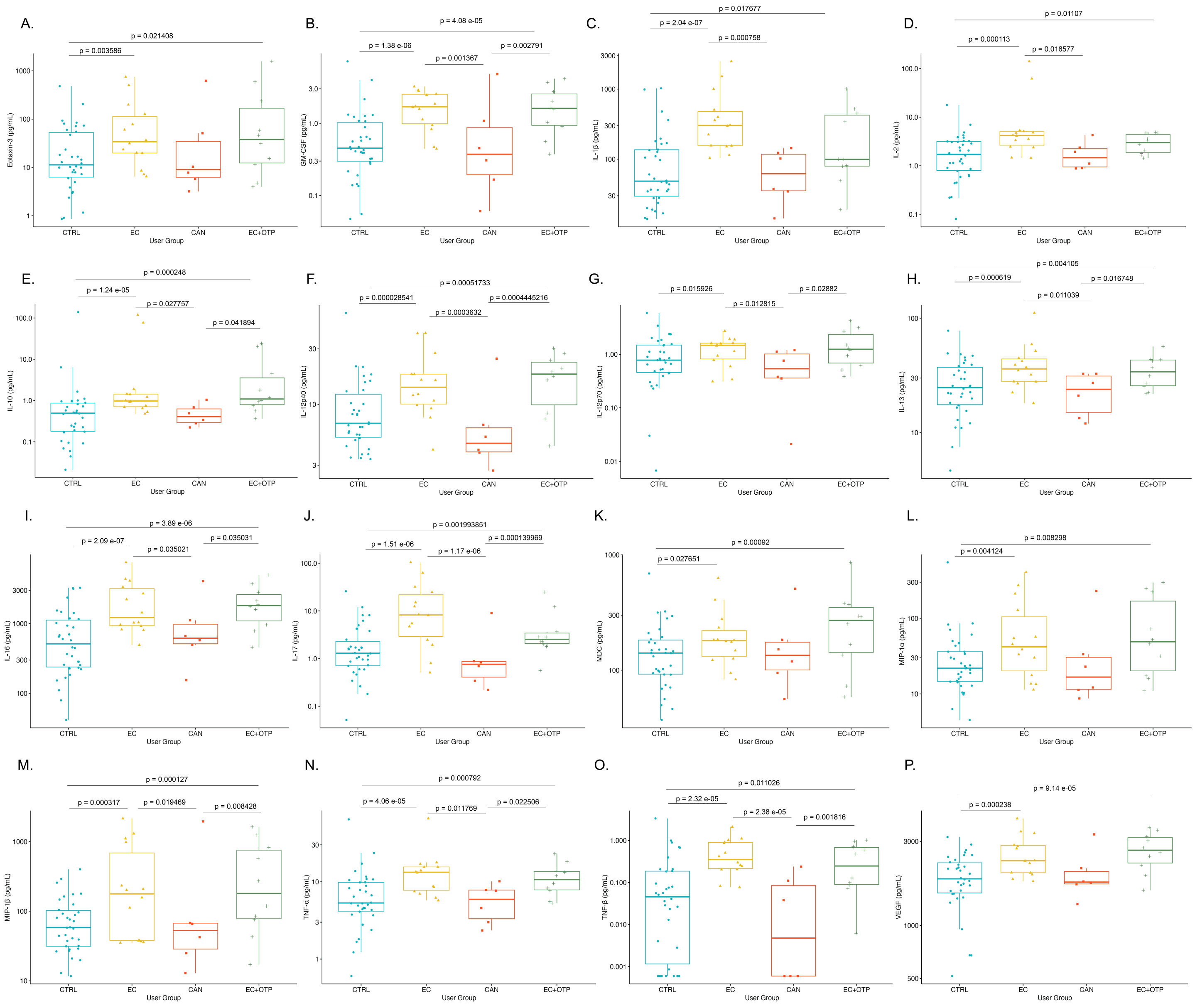

### Supplementary Figure 3

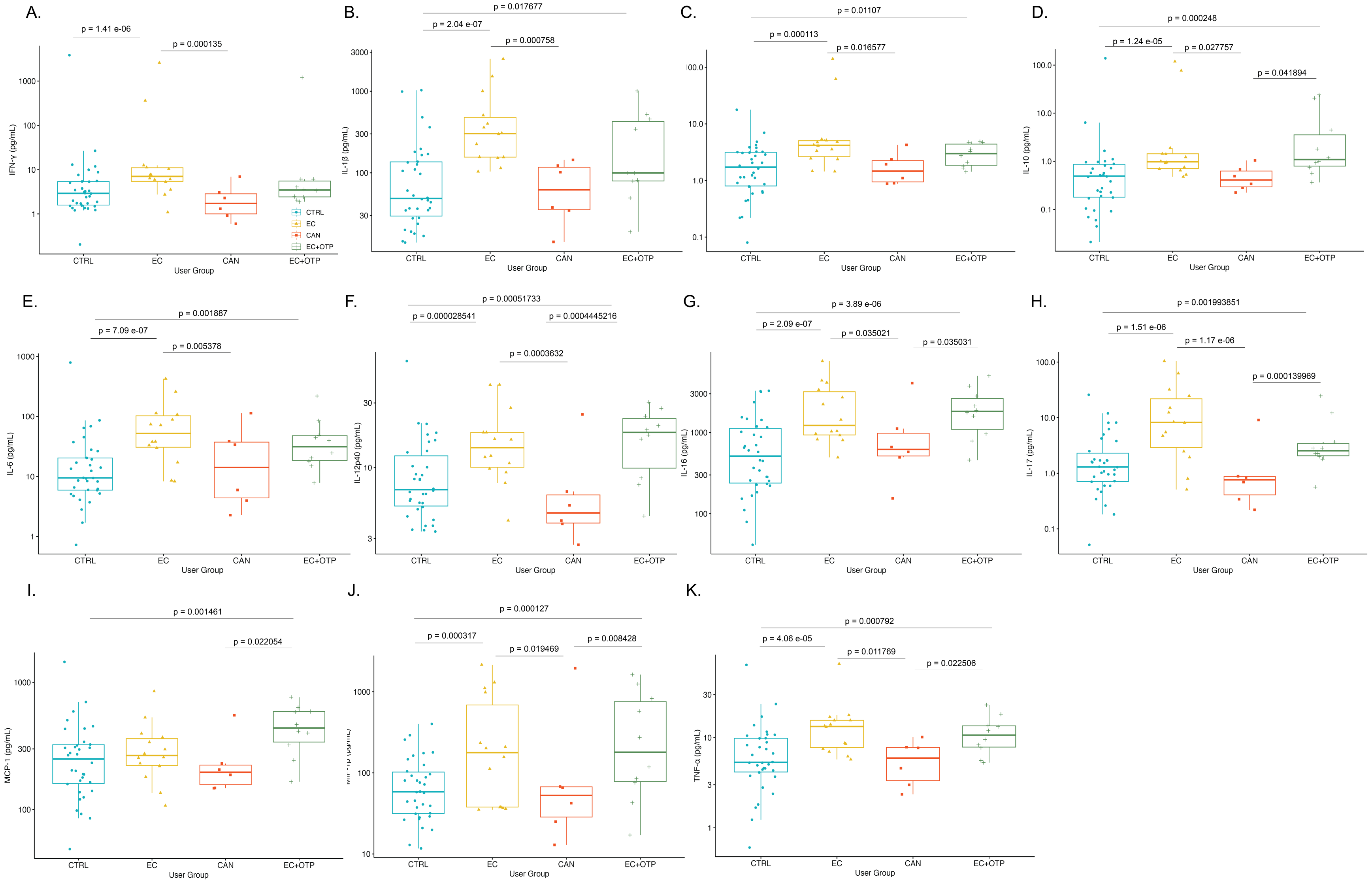

**Supplementary Figure 4**

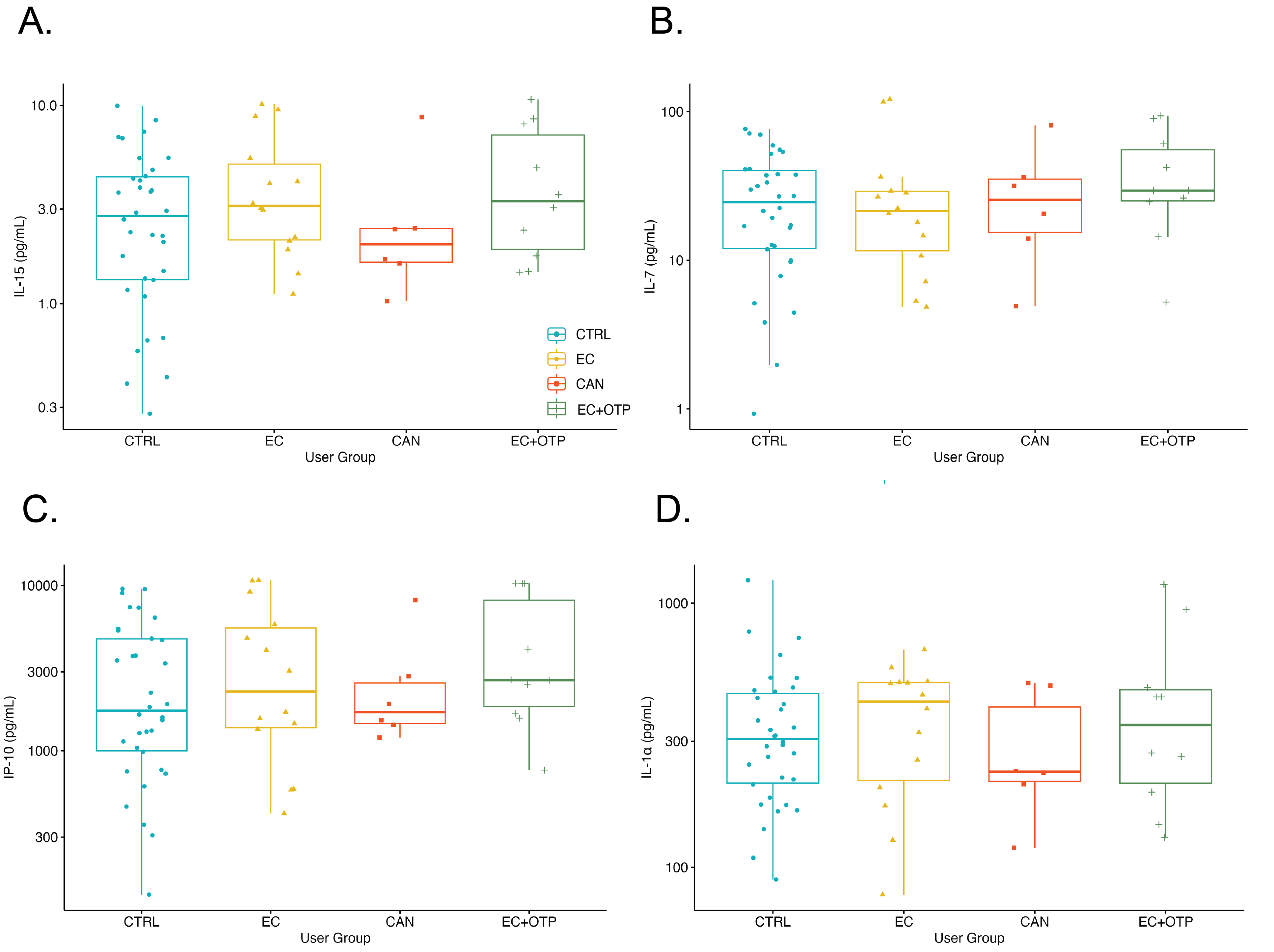

#### Supplementary Figure 5

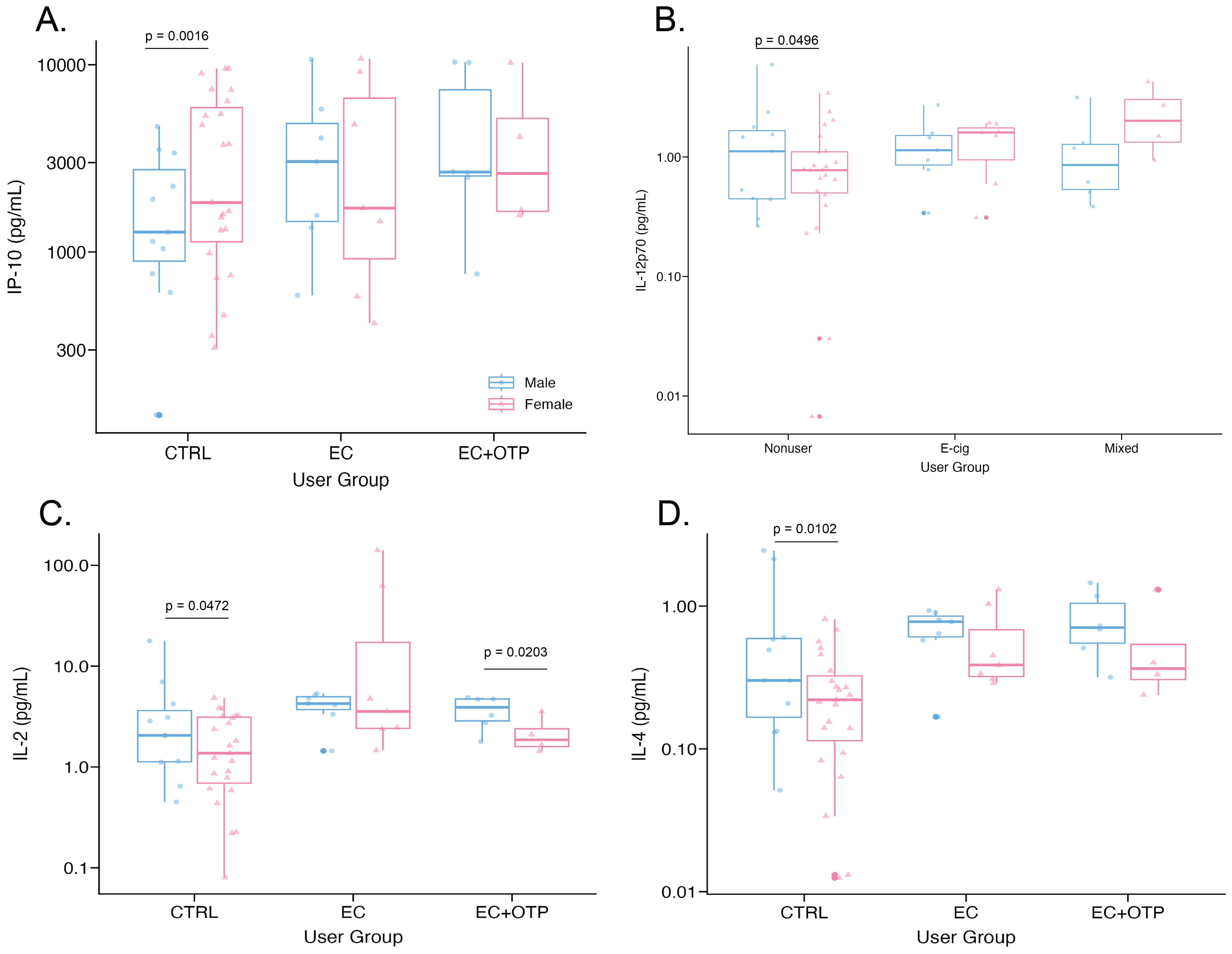

**Supplementary Figure 6**

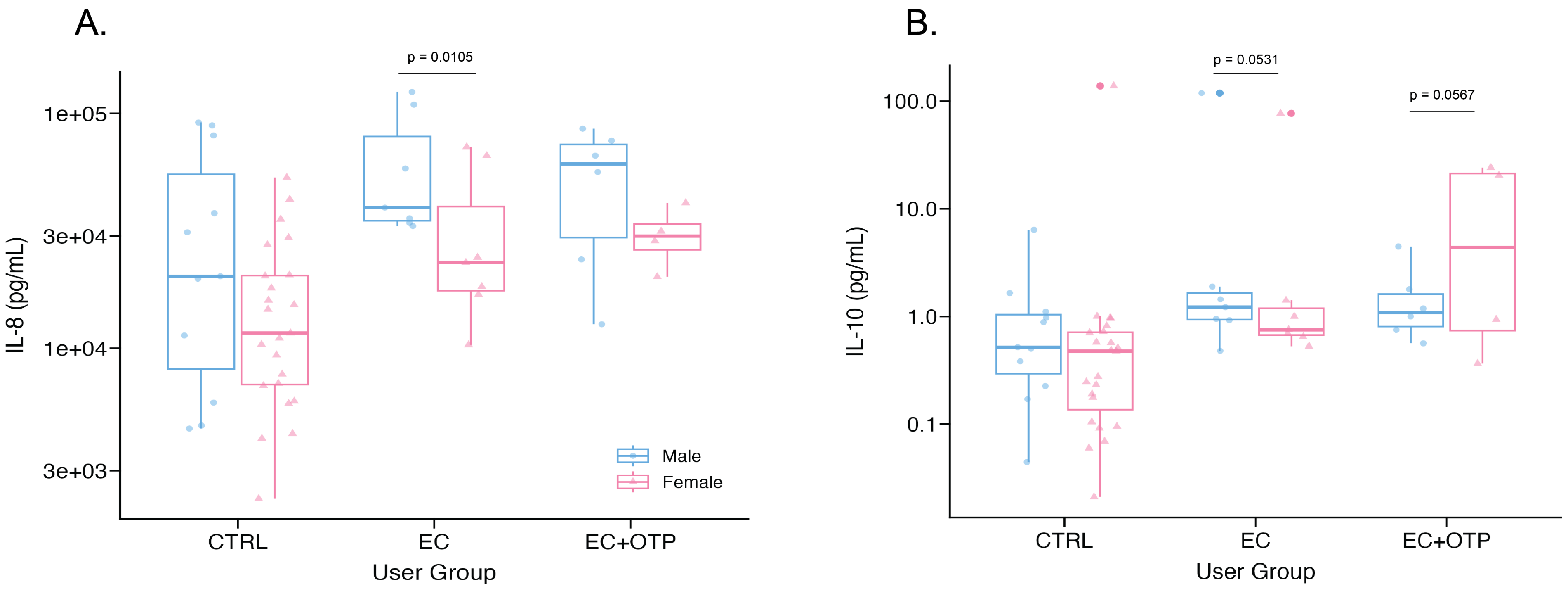

#### Supplementary Figure 7

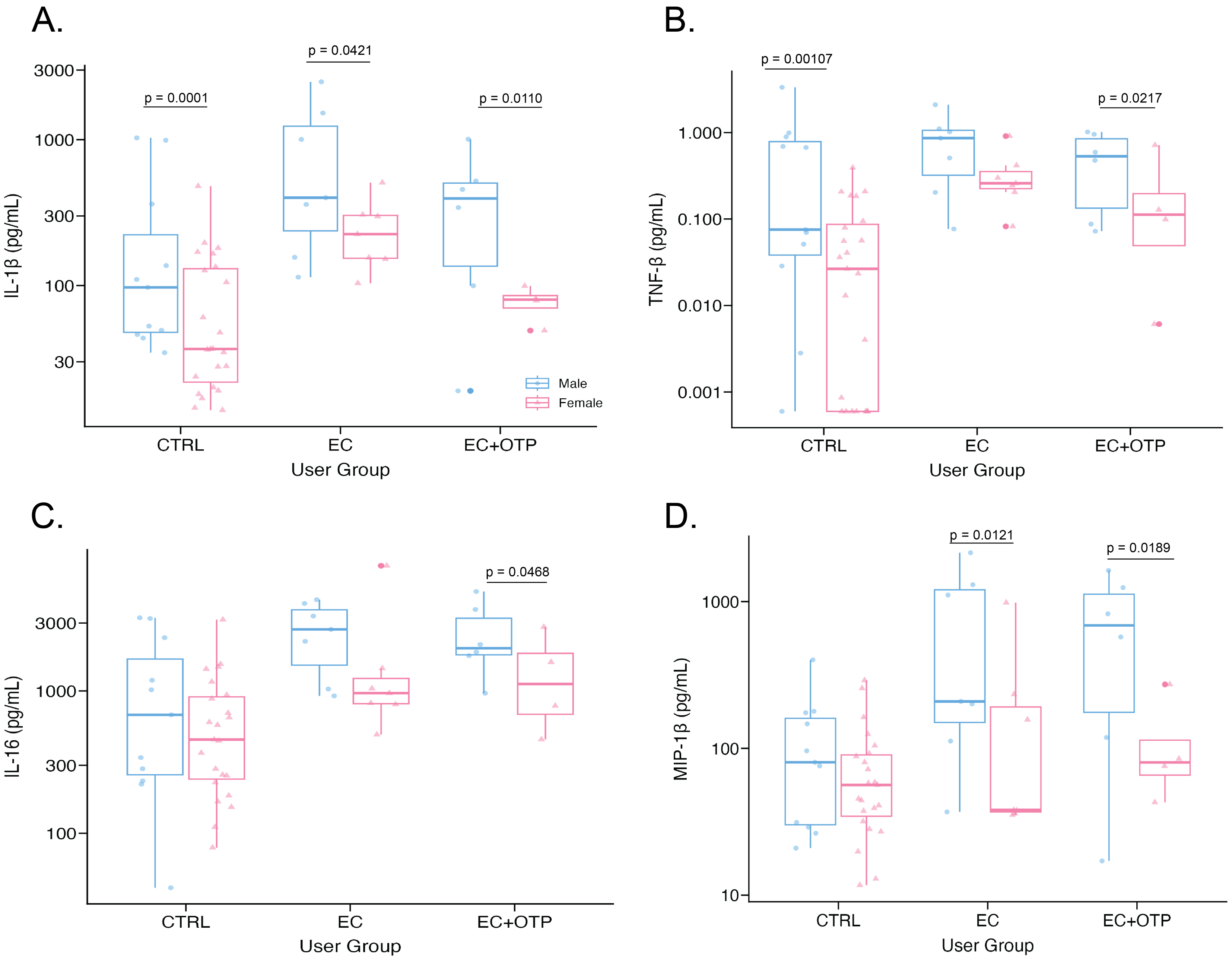

#### Supplementary Figure 8

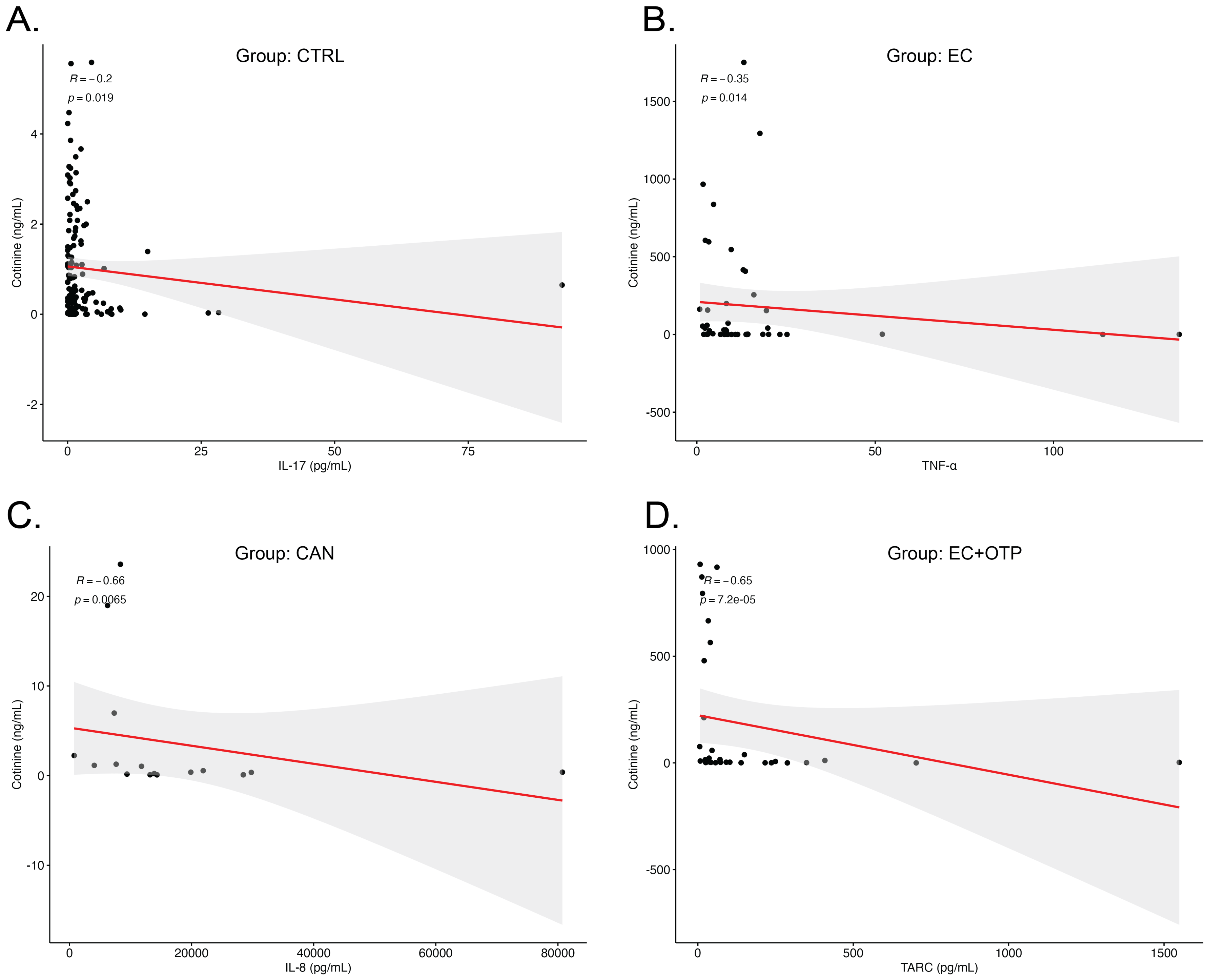
